# White matter microstructure in Parkinson’s disease with and without elevated REM sleep muscle tone

**DOI:** 10.1101/2021.10.26.21265550

**Authors:** Rémi Patriat, Pramod K. Pisharady, Sommer Amundsen-Huffmaster, Maria Linn-Evans, Michael Howell, Jae Woo Chung, Matthew N. Petrucci, Aleksandar Videnovic, Erin Holker, Joshua De Kam, Paul Tuite, Christophe Lenglet, Noam Harel, Colum D. MacKinnon

## Abstract

People with Parkinson’s disease who have elevated muscle activity during rapid eye movement sleep (REM sleep without atonia) typically have worse motor and cognitive impairment compared to those with normal muscle atonia during REM sleep. This study used tract-based spatial statistics to compare diffusion MRI measures of fractional anisotropy, radial, mean and axial diffusivity (measures of axonal microstructure based on the directionality of water diffusion) in white matter tracts between people with Parkinson’s disease with and without REM sleep without atonia and controls and their relationship to measures of motor and cognitive function. Thirty-eight individuals with mild-to-moderate Parkinson’s disease and twenty-one matched control subjects underwent ultra-high-field MRI (7Tesla), quantitative motor assessments of gait and bradykinesia, and neuropsychological testing. The Parkinson’s disease cohort was separated post-hoc into those with and without elevated chin or leg muscle activity during REM sleep based on polysomnography findings. Fractional anisotropy was significantly higher, and diffusivity significantly lower, in regions of the corpus callosum, projection, and association white matter pathways in the Parkinson’s group with normal REM sleep muscle tone compared to controls, and in a subset of pathways relative to the Parkinson’s disease group with REM sleep without atonia. The Parkinson’s disease group with elevated REM sleep muscle tone showed significant impairments in gait and upper arm speed compared to controls and significantly worse scores in specific cognitive domains (executive function, visuospatial memory) compared to the Parkinson’s disease group with normal REM sleep muscle tone. Regression analyses showed that gait speed and step length in the Parkinson’s disease cohort were predicted by measures of fractional anisotropy of the anterior corona radiata, whereas elbow flexion velocity was predicted by fractional anisotropy of the superior corona radiata. Visuospatial memory task performance was predicted by the radial diffusivity of the posterior corona radiata. These findings show that people with mild-to-moderate severity of Parkinson’s disease who have normal muscle tone during REM sleep demonstrate compensatory-like adaptations in axonal microstructure that are associated with preserved motor and cognitive function, but these adaptations are reduced or absent in those with increased REM sleep motor tone.

## Introduction

Under normal physiological circumstances, REM sleep is characterized by vivid dream mentation combined with skeletal muscles paralysis. In people with Parkinson’s disease, muscle activity during REM sleep is often abnormally elevated (termed REM sleep without atonia, RSWA) and when observed in conjunction with dream enactment (complex movements or vocalizations) reflects the co-expression of REM Sleep Behavior Disorder (RBD). ^1-8^ Isolated RBD is recognized as a prodromal stage of overt synucleinopathy with approximately 75% of individuals eventually phenoconverting to Parkinson’s disease, dementia with Lewy Bodies, or multiple system atrophy within 12 years.^9^ While isolated RBD is estimated to be present in about 1% of the population,^10^ approximately 30-50% of individuals with Parkinson’s disease meet clinical criteria for diagnosis of RBD based on polysomnography-derived evidence of RSWA and dream enactment behavior. The co-expression of Parkinson’s disease with RBD is associated with an increased likelihood and severity of cognitive impairment and a motor presentation that is dominated by an akinetic-rigid syndrome, postural instability, and gait disturbances compared to those without RBD.^11-15^ Currently, the mechanisms contributing to increased involvement of motor and cognitive systems in Parkinson’s disease with RBD are poorly understood.

Differences in the expression and rate of progression of Parkinson’s disease between individuals reflect, in part, the extent of neuronal loss in nigral and extranigral structures, but adaptive and maladaptive changes in the structural and functional connectivity of extranigral networks may also contribute to the masking and unmasking of motor and non-motor signs.^16^ Alterations in white matter microstructure, as assessed using measures derived from diffusion magnetic resonance imaging (dMRI), have been described in people with early-stage Parkinson’s disease.^17-19^ These studies have shown significantly higher fractional anisotropy (FA) and lower mean and radial diffusivities (MD, RD), reflecting increased directionality of water diffusion, in callosal, projection and association fibers in early-stage Parkinson’s disease compared with controls. Worsening of disease severity is associated with a progressive decrease in FA and increase in MD and RD.^19-27^ FA in the corpus callosum has also been shown to be inversely correlated with clinical measures (UPDRS III) of disease severity.^28^ The presence of increased FA across diffuse white matter tracts in early-stage Parkinson’s disease may reflect adaptive “neural compensations” possibly acting to preserve motor and cognitive function, or masking the expression of signs of early neurodegenerative changes in white matter, such as the loss of cortico-cortical and transcallosal fibers.^18, 28^ The latter can lead to increased anisotropy in regions where corticofugal and crossing fibers intersect.

This study used ultra-high field (7Telsa – 7T) MRI and tract-based spatial statistics (TBSS) to: (1) compare the microstructure of white matter pathways between people with Parkinson’s disease with (PD+RSWA) and without (PD-RSWA) elevated REM sleep muscle tone, and sex, age and education matched healthy adults, and (2) examine the relationships between measures of white matter microstructure and quantitative measures of motor and cognitive function in the Parkinson’s disease cohort. We hypothesized that measures of FA would be significantly higher in motor and prefrontal white matter tracts in the PD-RSWA compared to the PD+RSWA group and that the level of FA would be significantly correlated with quantitative measures of gait, arm speed and cognitive function.

In keeping with our hypotheses, the results showed that FA was significantly higher (RD and MD lower) in multiple regions of the white matter in the PD-RSWA group compared to matched healthy older adults and the PD+RSWA group. Based on our results and the previously reported finding that elevated white matter FA is a characteristic of early-stage disease,^28^ we hypothesize that the elevated white matter FA in the PD-RSWA group reflects adaptive compensatory neuroplastic changes in axonal microstructure, and the absence of elevated FA in the PD+RSWA cohort reflects increased severity of disease.

## Materials and methods

### Study participants and design

Sixty-two participants completed testing. Two participants were excluded post-hoc from the analysis due to image quality issues (e.g., excessive subject motion), and one participant was excluded due to a change in diagnosis. The final dataset included data from 38 individuals with mild-to-moderate severity of Parkinson’s disease and 21 healthy older adults who were sex- and age-matched to the Parkinson’s disease group. A priori sample size (minimum of 14/group) was determined based on an estimated effect size of 0.76 for the difference in mean FA of the corticospinal tract of controls and PD15 (power = 85%, alpha = 0.05). A summary of the demographics and clinical characteristics of the participants is shown in Table 1. Clinical diagnosis of Parkinson’s disease was determined by movement disorders neurologists according to the Movement Disorder Society clinical diagnostic criteria for Parkinson’s disease.^29^ Inclusion criteria included age 21-79 years and the ability to ambulate 50 m without an assistive device. Participants were excluded for any additional neurological disorder, a score of less than 22/30 on the Montreal Cognitive Assessment (MoCA),^30, 31^ musculoskeletal disorders that significantly affected movement, implanted deep brain stimulators or other surgeries to treat Parkinson’s disease, pregnancy, untreated sleep apnea, or the presence of non-MRI compatible medical implants or devices.

**Table 1.** Demographic and clinical characteristics of the participants.

|  | Healthy Older Adults | PD-RSWA | PD+RSWA | Significance (p) |
| --- | --- | --- | --- | --- |
| N (# of females)<sup>1</sup> | 21 (11) | 20 (6) | 18 (9) | 0.294 |
| Age (years)<sup>2</sup> | 61.3 ± 8.0 | 63.0 ± 8.6 | 65.8 ± 4.8 | 0.206 |
| Education (years)<sup>2</sup> | 16.0 ± 2.5 | 16.1 ± 1.0 | 16.8 ± 2.6 | 0.517 |
| MoCA scores<sup>3</sup> | 26.8 ± 2.0 | 27.6 ± 1.8 | 27.5 ± 1.9 | 0.358 |
| Years since diagnosis<sup>4</sup> | N/A | 2.1 ± 1.5 | 2.8 ± 2.2 | 0.079 |
| MDS-UPDRS III<sup>5</sup> | N/A | 34.4 ± 12.0 | 41.2 ± 14.5 | 0.118 |
| Hoehn and Yahr Stage<sup>4</sup> | N/A | 2.1 ± 0.3 | 2.3 ± 0.5 | 0.082 |
| Levodopa Eq (mg)<sup>4</sup> | N/A | 140 ± 175 | 365 ± 397 | **0.032** |

Statistical tests performed to compared groups: <sup>1</sup> Chi Square, <sup>2</sup> Kruskal-Wallis (H), <sup>3</sup> Univariate ANOVA, <sup>4</sup> Mann-Whitney U, <sup>5</sup> T-Test. N/A = not applicable. <sup>a</sup> Levodopa equivalent dosages were calculated using the conversion factors provided by Tomlinson et al. <sup>76</sup>. PD+RSWA= Parkinson's disease with RSWA. PD-RSWA= Parkinson's disease without RSWA.

All participants, including control subjects, underwent a battery of clinical assessments including the Movement Disorders Society Unified Parkinson’s Disease Rating Scale (MDS-UPDRS) Part III evaluation, Hoehn and Yahr staging, a full neuropsychological assessment battery, overnight in-laboratory polysomnography (PSG), a 7T MRI scan and quantitative motor assessments. Motor testing, clinical examinations and MRI scans were conducted in the practically-defined off-medication state following overnight withdrawal of levodopa and dopamine agonists and/or 48-hour withdrawal from long-acting dopaminergic medications. Overnight PSG and neuropsychological testing were conducted with the participant in their usual medication state. All assessors and signal processors were blinded to the disease state and group category. The study was approved by the Institutional Review Board at the University of Minnesota, and written informed consent was obtained from all the participants according to the Declaration of Helsinki prior to inclusion into the study.

### Classification of participants according to RSWA

Overnight sleep studies were performed using standard video-based PSG procedures, including electromyography (EMG) recordings from the submentalis (chin), flexor digitorum superficialis and tibialis anterior (TA) muscles as well as electroencephalography recordings from 10 scalp electrodes. PSGs were scored by a single rater (AV), blinded to disease state, using the American Academy of Sleep Medicine Manual for the Scoring of Sleep and Associated Events to identify excessive sustained (tonic) and transient (phasic) muscle activity during REM sleep.^32^ The primary outcomes measures were the percentage of REM sleep with tonic or phasic EMG activity in the submentalis muscle and phasic activity in tibialis anterior muscle. The level of tonic or phasic RSWA was used to assign the Parkinson’s disease participants to a group with normal (PD-RSWA) or abnormally elevated (PD+RSWA) muscle tone. The presence/absence of REM sleep dream enactment behavior was not used in the group selection criteria due to concerns about potential false negative reporting from individuals who do not have a bed partner and variability in dream enactment behavior across PSG sessions.^33^ See Supplement 1 for more details about RSWA group classification procedures. Eighteen of the 38 participants with Parkinson’s disease were assigned to the PD+RSWA group. All participants in the PD+RSWA group had elevated phasic or tonic submentalis EMG and 14 of 18 (78%) had self-reported or partner-reported dream enactment and/or dream enactment behavior that was observed during the PSG.

### Motor testing

Quantitative measures of steady-state gait and ballistic arm movements were collected in all subjects. Spatial and temporal metrics of steady-state gait were collected by walking continuously in an oval-shaped course that contained a 10 m long pressure-sensitive gait mat (Gaitrite, Franklin, NJ, USA). A minimum of 35 steps were collected on the mat to ensure reliability.^34^ The primary outcome measures were gait speed, stride length, and right and left step length (normalized to leg length). Upper limb bradykinesia was assessed by having subjects perform a self-initiated (uncued) ballistic elbow flexion movement to a target at 72 degrees (± 3 deg. target zone) using a custom-built manipulandum. Subjects were instructed to move as “fast as possible to the target” and hold the final position until instructed to return to the start position. The primary outcome variable was peak velocity. The ballistic movement data were collected using a CED Power 1401 data acquisition interface paired with Signal software (Cambridge Electronic Design, Ltd., Cambridge, UK) at a sampling rate of 2000 Hz and processed using custom MATLAB software (MathWorks, Natick, MA, USA).

### Neuropsychological evaluations

The neuropsychological battery was designed to assess attention and executive functioning, initiation-perseveration, construction, conceptualization, memory, visuospatial abilities and visual memory, verbal memory, language abilities, motivation, mood state and emotion in people with Parkinson’s disease. These tests included: Spatial Span from Wechsler Memory Scale - 3), Wechsler Adult Intelligence Scale - IV (WAIS-IV; Matrix Reasoning, Digit Span), Rey Complex Figure (copy trial), Boston Naming Test – 2, Delis-Kaplan Executive Function System (D-KEFS Verbal Fluency), Stroop (Golden version), Wisconsin Card Sorting Test (WCST)- 64, Hopkins Verbal Learning Test – Revised, Brief Visuospatial Memory Test (BVMT) – Revised, Mattis Dementia Rating Scale 2 (DRS-2), Beck Depression Inventory-2 (BDI-2).

### MRI scanning protocol

Participants were scanned on a 7T MRI scanner (Magnetom 7T Siemens, Erlangen, Germany) at the Center for Magnetic Resonance Research of the University of Minnesota. The scanner was equipped with SC72 gradients capable of 70 mT/m and a 200 T/m/s slew rate using a 32-element head array coil (Nova Medical, Inc., Burlington, MA, USA). Whenever subject head size enabled enough space in the coil, dielectric pads were utilized in order to enhance signal in the temporal regions. The scanning protocol consisted of diffusion-weighted images covering the whole brain (50 gradient directions, b-value = 1500 s/mm^2^, 4 additional b0-volumes, 1.25 mm isotropic, 6.5 minutes scan time) and a T1-weighted image (0.6 mm isotropic, 6.5 minutes scan time).^35^ Two sets of diffusion data were collected during each session, with reversed phase encoding directions (anterior-to-posterior and posterior-to-anterior). These datasets were subsequently combined to correct for distortions and to increase the signal to noise ratio, as described below.

### MRI data processing and analysis

Fig. 1 shows a block diagram of the imaging data processing and analysis steps. The data were corrected for distortions due to eddy currents, susceptibility-induced off-resonance artifacts and subject motion.^36, 37^ A DTI model was subsequently fitted to the corrected data using FSL,^38^ and FA, RD, MD, and axial diffusivity (AD) were calculated. For the whole brain data analysis, we ran the TBSS algorithm in FSL, improved with tensor-based registration ^39^ and spatial normalization available in DTI-TK.^40^ The improved spatial normalization and tensor-based registration using DTI-TK has been shown to produce fewer false positives compared to the standard registration available in FSL.^40^ Once the data are registered, the TBSS performs a skeletonization of the white matter and projection of the DTI features, such as FA, onto this white matter skeleton. We conducted the TBSS analysis with four categories of pairings of the data: i) all Parkinson’s disease vs. healthy subjects, ii) PD-RSWA vs. healthy subjects, iii) PD+RSWA vs. healthy subjects, and iv) PD-RSWA vs. PD+RSWA.

**Figure 1.**
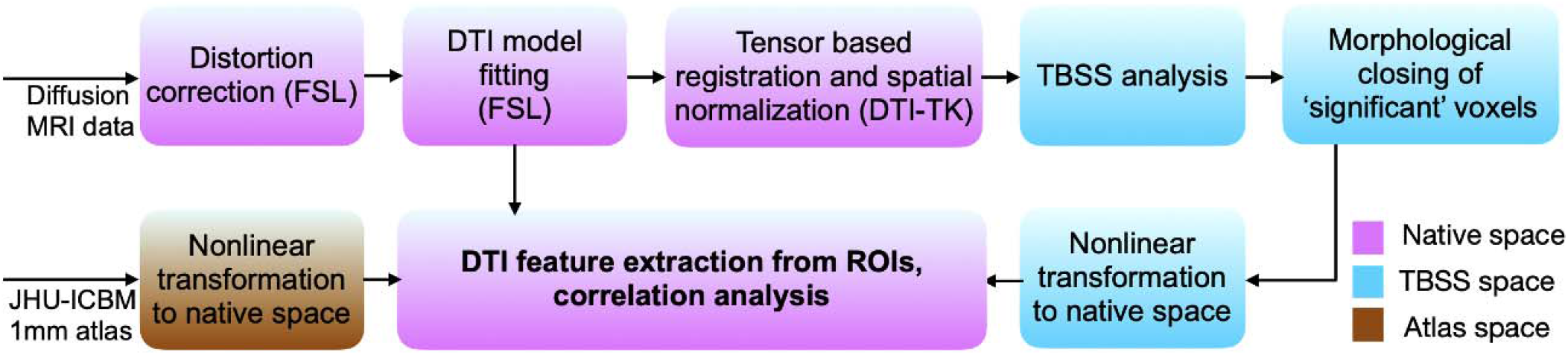
Flow diagram depicting the image processing and analysis pipeline. Words in parenthesis indicate the software being used for a particular step. TBSS = Tract-based spatial statistics. DTI = Diffusion tensor imaging. MRI = Magnetic resonance imaging.

### Statistical Analysis of MRI data

Nonparametric permutation inference of the skeletonized white matter was done using the ‘randomise’ algorithm ^41^ in FSL. The algorithm provides p-values corrected for multiple comparisons along the white matter skeleton, estimated using a threshold-free cluster enhancement. In order to extract diffusion features for each participant, a binary image of the significant clusters (from the group analysis) was first created. Then, a morphological closing operation with a kernel size of 3×3 voxels was performed to fill in any holes within the binary image. This closed binary image was transformed from the TBSS space to individual native space through linear and nonlinear registrations (‘flirt’ and ‘fnirt’ algorithms from FSL). The anatomical locations of the significant clusters were identified (for each subject) by computing the intersection of these regions with the JHU ICBM 1mm atlas ^41^ warped into the native data space. We then extracted the average of the diffusion features (FA, MD, RD, and AD) from the voxels within each of these intersecting regions of interest (ROIs).

### Statistical analysis of behavioral differences and associations with diffusion metrics

Demographic features between groups were compared using a one-way ANOVA (MoCA), t-test (MDS-UPDRS Part III total score), Mann-Whitney U (disease duration, Hoehn and Yahr stage, levodopa daily equivalent) or Kruskal-Wallis test (age, years of education). Homogeneity of variance of the behavioral variables was tested using Levene’s Test of Equality of Error Variances and either a univariate ANOVA or Kruskal-Wallis test was used to test for main effects of groups (PD+RSWA, PD-RSWA, healthy older adults). For variables with bilateral measures, a repeated measures ANOVA was used with factors of group and side. Age and sex were included as covariates in the analysis of the movement data and years of education was added as a covariate to the analyses of the cognitive data. Post-hoc interaction effects were modeled using Tukey’s Honestly Significant Difference test.

Partial correlations, controlled for age and sex, were used to examine associations between behavioral and white matter diffusion measures in the Parkinson’s disease cohort. A multivariate step-wise linear regression was used to identify the diffusion measures that best predicted the motor and cognitive outcomes measures. The regression analyses were controlled for age, sex, disease duration and levodopa equivalent daily dose for predictions of motor outcomes. Years of education was added as a factor for predictions of cognitive outcomes. Statistical analysis was performed using IBM SPSS 25 (IBM Corp., Armonk, NY, USA) and the significance level was set at p < 0.05 for the ANOVA/Kruskal-Wallis and post hoc tests. The statistical threshold for the correlation and multivariate regression analyses was set at p < 0.01.

### Data availability

The derived data that support the findings of this study will be made available on reasonable request, for the purposes of reproducing the results presented, subject to institutional approval.

## Results

### Demographics

There were no significant differences between the control, PD-RSWA and PD+RSWA groups for the variables sex, age, education and MoCA score (Table 1). Similarly, there was no significant difference between the Parkinson’s disease groups in age at diagnosis, years since diagnosis, Hoehn and Yahr stage or total MDS UDRS III score. Levodopa equivalent dose was significantly higher in the PD+RSWA group compared with the PD-RSWA group (p = 0.032).

### TBSS: Parkinson’s disease versus healthy older adults

Higher FA and lower MD, RD, and AD were observed in the Parkinson’s disease compared to the control group (Fig. 2). MD and RD presented more spatially widespread differences between groups than FA and AD. Clusters with significantly higher FA or lower RD and MD in the Parkinson’s disease group included: corpus callosum (regions of genu, body and splenium), anterior and posterior limbs of the internal capsule, external capsule, anterior, superior and posterior corona radiata, posterior thalamic radiation, superior longitudinal fasciculus, superior fronto-occipital fasciculus, tapetum, and cerebral peduncle (bilateral findings for all the ROIs). Lower MD and RD were also seen in the cingulum and sagittal stratum in the Parkinson’s disease group. Lower AD was observed in the corpus callosum (regions of genu and body), anterior limb of internal capsule (only left), external capsule (only left), and anterior and superior corona radiata (both bilateral).

**Figure 2.**
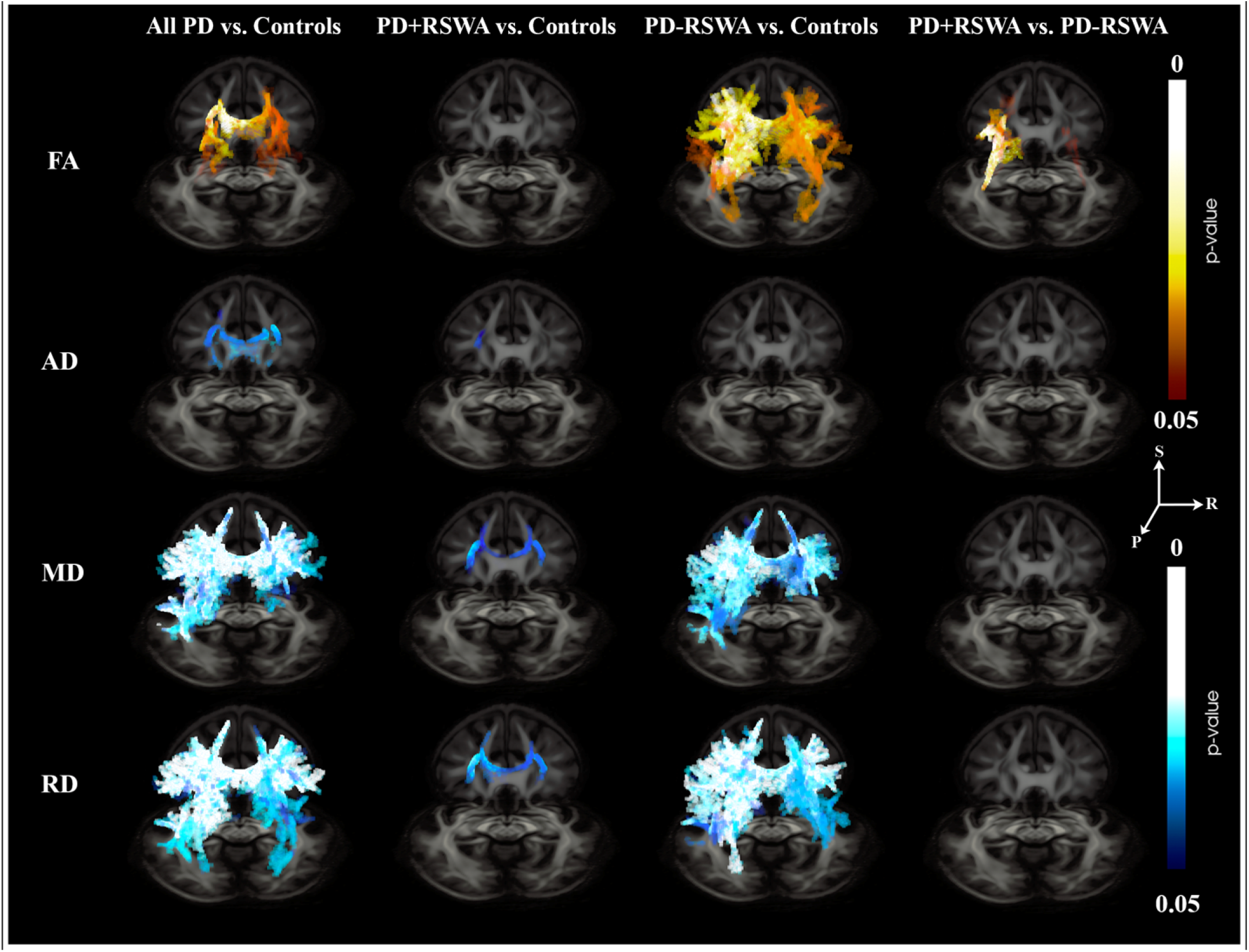
3D axial views TBSS results showing the regions where statistically significant differences between different groups (columns) were observed, in different diffusion metrics (rows). S= superior, R = right, P = posterior. Warm colors show a positive difference while cold colors show a negative difference.

**Figure 3.**
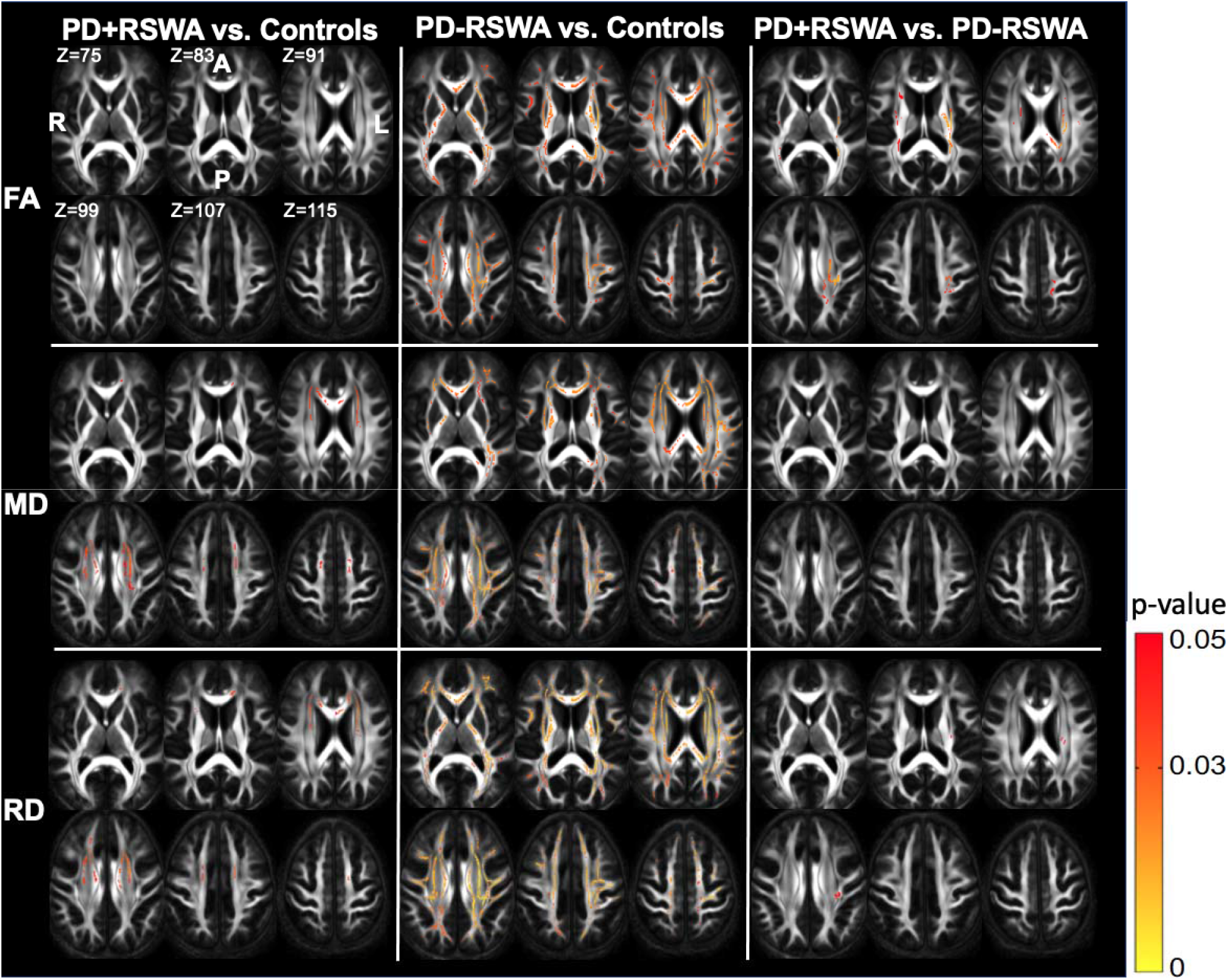
2D axial views of the TBSS results showing the main regions where statistically significant differences between different groups (columns) were observed, in different diffusion metrics (rows). R = right, L = left, A = anterior, P = posterior.

### TBSS: PD+RSWA versus healthy older adults

MD and RD were significantly lower in the PD+RSWA group in the corpus callosum (regions of genu and body), anterior limb of the internal capsule, external capsule, and anterior and superior corona radiata (bilateral findings for all the ROIs, Fig. 2 & 3). Lower AD in the PD+RSWA group was seen in the anterior and superior corona radiata (only left side for both ROIs).

### TBSS: PD-RSWA versus healthy older adults

The PD-RSWA group exhibited significantly higher FA values and lower MD and RD compared to the healthy control group in many regions including the corpus callosum (regions of genu, body and splenium), anterior and posterior limbs of the internal capsule, retrolenticular part of internal capsule, external capsule, anterior, superior and posterior corona radiata, posterior thalamic radiation, superior longitudinal fasciculus, superior fronto-occipital fasciculus, tapetum, cingulum (bilateral findings for all the ROIs), and sagittal stratum (only left, Fig. 2 & 3). In addition, higher FA was observed in the regions of cerebral peduncle (bilateral), corticospinal tract (only right), and superior cerebellar peduncle (only right). AD did not show significant differences in any of these regions.

### TBSS: PD+RSWA versus PD-RSWA

The PD-RSWA group showed higher FA compared to the PD+RSWA group in multiple regions including corpus callosum (regions of body and splenium), anterior and posterior limbs of the internal capsule, retrolenticular part of internal capsule, external capsule, anterior, superior, and posterior corona radiata, posterior thalamic radiation, superior longitudinal fasciculus, superior fronto-occipital fasciculus, and tapetum (bilateral for all the ROIs) (Fig. 2 & 3). Lower RD was seen in the PD-RSWA group in the regions of posterior limb of internal capsule, superior and posterior corona radiata, external capsule, and superior longitudinal fasciculus (only left for all the ROIs), compared to the PD+RSWA group. There were no significant differences in MD and AD between these groups.

### Motor Assessments

Significant main effects of group were seen for measures of normalized gait speed, stride, and step lengths (F > 3.612, p < 0.035; no effect of side or side x group interaction for step length) (Fig. 4). Sex, but not age, was a significant covariate for gait speed (p = 0.025). Post-hoc analyses demonstrated that gait speed, stride, and step lengths (bilateral) were significantly lower in the PD+RSWA group compared with healthy older adults (p < 0.037). There was no significant group effect for cadence and no significant differences in gait measures between the PD+RSWA and PD-RSWA groups. Peak elbow flexion velocity during ballistic arm movement also showed a significant main effect of group (F = 3.747, p = 0.031). Age and sex were significant covariates (p < 0.018). There was no effect of side or side x group interaction effect. Post-hoc tests showed that movement speed was significantly lower in the PD+RSWA group compared with healthy older adults for both right and left arm movements (p < 0.034).

**Figure 4.**
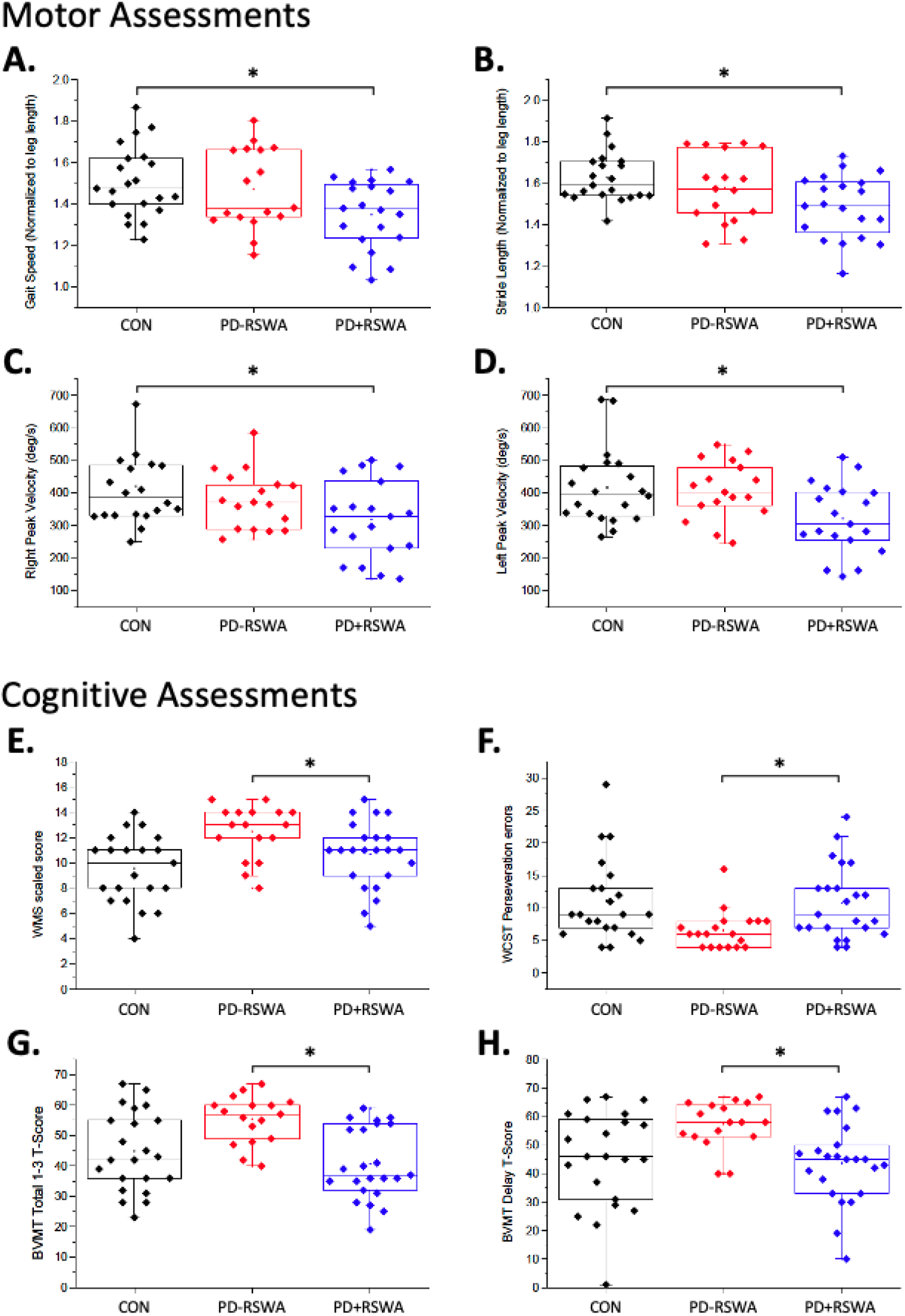
Summary of behavioral results. Plots A-B show the average normalized gait speed and normalized stride length during steady-state gait for each individual in the control (CON=black), PD-RSWA (red) and PD+RSWA (blue) groups. Plots C-D show the average peak velocities during ballistic elbow movement. Normalized gait speed, step length and right and left peak elbow movement velocity were significant lower in the PD+RSWA group compared to healthy older adults (^*^ = p < 0.037). Plots E-H show the results from some of the cognitive tests including the Wechsler Memory Scale – Scaled Score (WMS) (plot E), Wisconsin Card Sorting Task (WCST) – total number of perseveration errors (plot F), Brief Visual Memory test (BVMT) – total score from trials 1 to 3 (plot G), and BVMT delay T-score (plot G). Scores for each of these tests were significantly lower in the PD+RSWA group compared to the PD-RSWA group (^*^ = p < 0.042). PD+RSWA= Parkinson’s disease with RSWA. PD-RSWA= Parkinson’s disease without RSWA. A univariate ANOVA or Kruskal-Wallis test was used to test for main effects of group (PD+RSWA, PD-RSWA, healthy older adults) with age, sex and years of education (for cognitive variables) as covariates.”

### Neuropsychological Assessments

Cognitive test performance was different between the PD-RSWA and PD+RSWA groups in specific domains (Fig. 4). There were main effects of group for measures of spatial-sequential working memory (Spatial Span subtest of the WMS-3 : F > 6.16, p < 0.005), visuospatial memory (BVMT-R, Trials 1, 2, 3 and total, delay raw score and T-score: F > 6.795, p < 0.003) and executive function (WCST-64, number of categories, perseveration errors, conceptualization level response: F > 5.05, p < 0.011). Post-hoc tests showed that spatial-sequential working memory scores and executive function scores were significantly better in the PD-RSWA compared to the PD+RSWA group (p < 0.042). Visuospatial memory was significantly better in the PD-RSWA group compared to both the PD+RSWA group and healthy older adults (p < 0.04). No significant differences between groups were seen for the other neuropsychological tests.

### Relationships between white matter diffusion measures and behavior in participants with PD

Less severity of motor disease and better performance on motor tasks was associated with higher FA (or lower RD and MD) (Table 2). Total motor MDS-UPDRS III and axial subscore were inversely correlated with FA in the right superior corona radiata. The MDS-UDPRS III axial subscore was positively correlated with RD in the right superior corona radiata, right and left superior longitudinal fasciculus and MD in the left superior longitudinal fasciculus. Step-wise linear regression showed that total MDS-UPDRS III score was predicted by FA of the right superior corona radiata whereas MDS-UPDRS axial subscore was predicted by RD of the right superior longitudinal fasciculus.

**Table 2.**
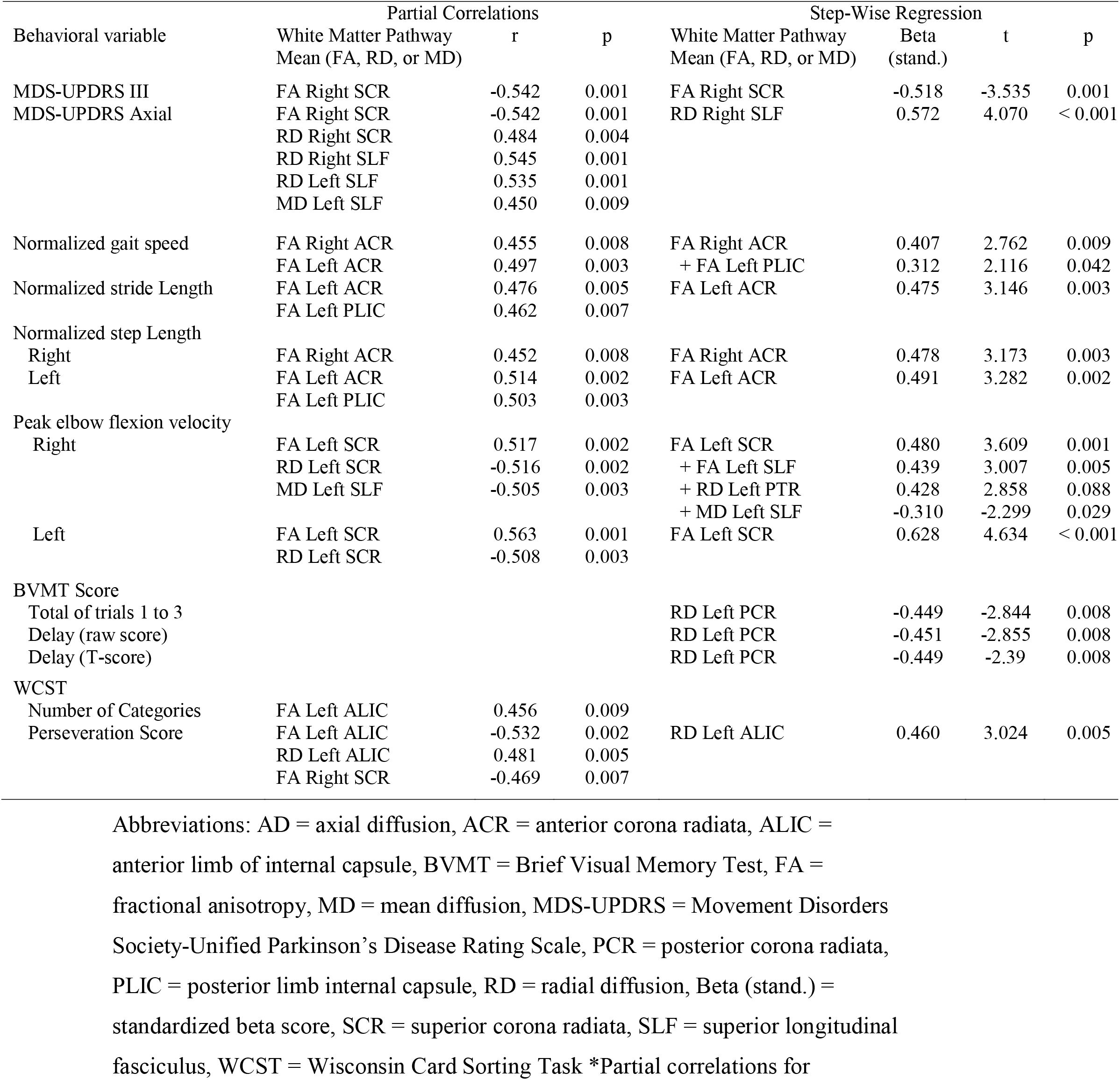
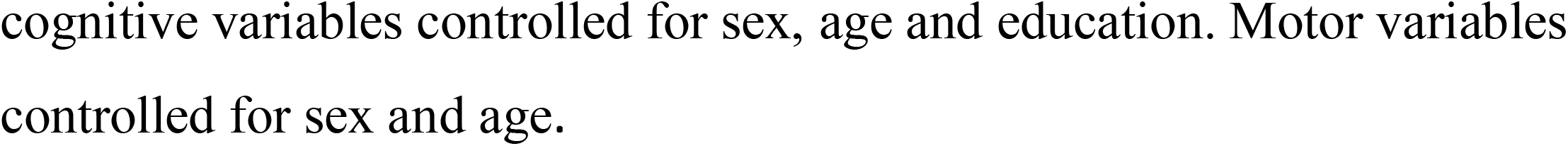
Summary of significant partial correlations and regressions between motor and cognitive outcome measures and mean white matter pathway diffusion measures.

Insert Table 2 Near Here

Normalized gait speed positively correlated with FA of the right and left anterior corona radiata (Fig. 5). Normalized stride length correlated with FA of the left anterior corona radiata and left posterior limb of the internal capsule. Normalized step length (right) correlated with FA of the right anterior corona radiata while the left side correlated with FA of left anterior corona radiata and left posterior limb of the internal capsule. Step-wise linear regression analyses showed that a model that included FA of the right anterior corona radiata and left posterior limb of the internal capsule significantly predicted normalized gait speed. FA of the left anterior corona radiata significantly predicted normalized stride length and left step length, whereas FA of the right anterior corona radiata predicted right step length. Right and left peak elbow flexion velocity were positively correlated with FA of the left superior corona radiata and inversely correlated with RD of the left superior corona radiata. Right elbow flexion velocity was also inversely correlated with MD of the left superior longitudinal fasciculus. Multiple regression analyses showed that right peak elbow flexion velocity was significantly predicted by FA of the left superior corona radiata and superior longitudinal fasciculus, RD of posterior thalamic radiation and MD of superior longitudinal fasciculus. Left peak elbow flexion velocity was significantly predicted by a model that included only FA of the left superior corona radiata (right superior corona radiata FA also predicted left velocity, p < 0.003, but did not add further to the fit of the model beyond that provided by the left superior corona radiata).

**Figure 5.**
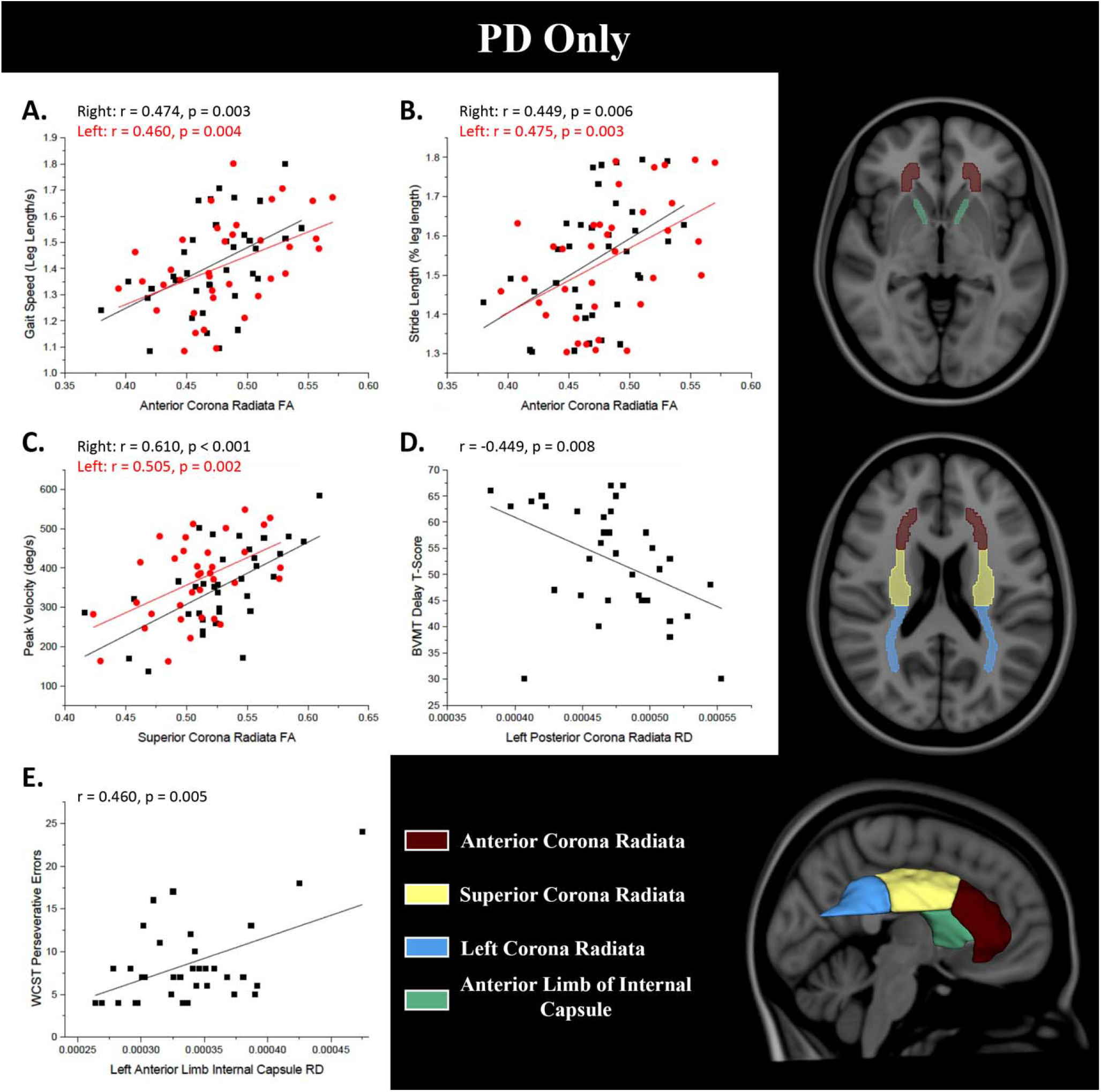
Pearson’s correlations between diffusion measures and behavior: (A) anterior corona radiata FA vs. stride length, (B) anterior corona radiata FA vs. gait speed, (C) superior corona radiata FA vs. peak elbow movement velocity, (D) left posterior corona radiata RD vs. Brief Visual Memory Test (BVMT) delay T-score, (E) left anterior limb of the internal capsule RD vs. Wisconsin Card Sorting Task (WCST) total number of perseveration errors. Hemis. = hemisphere, L = left, R = right.

Better performance on the neuropsychological assessments in the Parkinson’s disease group was associated with higher FA or lower RD, MD, and AD in specific white matter pathways. Partial correlations between cognitive and diffusion measures did not reach significance after control for age, sex, and education. However, regression analyses showed that RD of the left posterior corona radiata was a significant predictor of the BVMT (including scores for trials 1, 2 and 3, total of trials 1-3, total of trial 1-3 T-score and the delay raw and delay T-scores (Table 2). The perseveration score for the WCST was predicted by RD of the anterior limb of the internal capsule.

Wisconsin Card Sorting Task (WCST) total number of perseveration errors. Hemis. = hemisphere, L = left, R = right.

## Discussion

There were two main findings from this study: First, FA was significantly higher (RD and MD lower) in multiple regions of the white matter in the participants with Parkinson’s disease who had normal REM sleep muscle activity (PD-RSWA), compared to individuals with abnormally elevated REM motor tone (PD+RSWA) and matched healthy older adults. Second, higher FA (or lower MD, RD) in specific pathways was associated with less motor impairment and better cognitive function in the Parkinson’s disease cohort.

Our finding of higher FA (lower RD, MD) in the Parkinson’s disease group with normal REM sleep atonia (PD-RSWA) relative to healthy older adults is consistent with many previous studies showing diffuse white matter differences in individuals with early-stage Parkinson’s disease.^17, 19, 28, 42, 43^ These papers reported significant differences in many of the pathways identified in the present study including the corpus callosum, anterior and superior corona radiata, anterior and posterior internal capsule, thalamic radiation and superior longitudinal fasciculus. Wen et al. ^28^ further showed that FA was significantly more elevated in individuals with Hoehn and Yahr Stage 1 Parkinson’s disease compared to those with Hoehn and Yahr Stage 2 disease and that UPDRS III motor scores were inversely correlated with white matter FA and positively correlated with RD and MD. Importantly, prospective studies that have assessed changes in diffusion measures over the course of a year or more have shown a decrease in white matter FA or increase in RD and MD in association with increasing severity of disease.^19-27^ Taken together, these findings suggest that the course of disease may be characterized, in some individuals, by an early period during which white matter FA is higher (lower RD and MD) than matched healthy older adults, particularly in callosal (genu, body), projection (anterior and superior corona radiata, internal capsule), and association (superior longitudinal fasciculus, posterior thalamic radiation) pathways, followed by a progressive decline in FA, in concert with worsening of motor disease severity, to levels at or below those seen in healthy older adults.

The novel finding of the present study was the observation that, when a cohort with mild-to-moderate severity of disease was separated, post hoc, into groups with normal (PD-RSWA) and abnormally elevated REM sleep motor tone (PD+RSWA), only the PD-RSWA group showed significantly higher white matter FA relative to healthy older adults. The PD-RSWA group also showed significantly lower RD and MD across extensive white matter areas, compared to the healthy older adults. Conversely, no significant differences in FA were observed between the PD+RSWA group and controls, and differences in MD, RD and AD were restricted to a small subset of tracts. The absence of a significant difference in FA in the PD+RSWA group could be explained by: (1) an earlier onset or increased rate of disease, such that neuroplastic adaptations in white matter microstructure occur predominantly during the prodromal phase, or (2) phenotypic differences in axonal microstructure and/or the capacity for neuroplasticity relative to those without RSWA.

If elevated white matter FA is a characteristic of early-stage disease ^28^ and progressively decreasing FA (from elevated, to normal, to below normal) reflects disease progression,^27^ it could be argued that the absence of significantly elevated FA in the PD+RSWA cohort reflects increased severity of disease, possibility due to an earlier onset and/or faster rate of progression in both motor and non-motor circuits relative to the participants without RSWA. This idea is corroborated by work showing that white matter FA in the corticospinal tract, corpus callosum and superior longitudinal fasciculus is significantly increased in people with isolated RBD compared to controls,^44^ suggesting that elevated FA emerges during the prodromal phase of disease and that, by the time of diagnosis, white matter microstructural alterations have evolved to the point where there are no significant differences in FA from controls and may continue to progress to levels below controls with disease progression. While there were no significant differences in clinical scores between the Parkinson’s disease groups, quantitative motor assessments showed that gait and arm movement velocity were significantly impaired in the PD+RSWA group relative to the healthy older adult group, but not in the PD-RSWA group. Frontal executive function and visuospatial memory were also significantly more impaired in the PD+RSWA compared to the PD-RSWA group, which is consistent with studies showing greater cognitive impairment in people with Parkinson’s disease with RBD compared to those without RBD.^14, 45, 46^ Longitudinal studies have also provided evidence that the rate of decline of cognitive and motor symptoms is greater in people with Parkinson’s disease and RBD compared to those without RBD dopaminergic denervation.^47, 48^ The extent to which differences in motor and cognitive between people with Parkinson’s disease with and without RBD reflect the state of neurodegeneration of nigrostriatal dopaminergic neurons is currently unclear. Cross-sectional studies comparing groups with Parkinson’s disease with and without RBD (or probable RBD) using radiotracer assays of nigrostriatal dopaminergic function have been inconsistent, some showing significant reductions in people with RBD ^49, 50^ while others found no differences between groups.^51, 52^ Nonetheless, the balance of evidence demonstrates that the severity of disease, as reflected in clinical and quantitative measures of motor and cognitive function, is greater in people that co-express Parkinson’s disease and RBD, when compared to a cohort matched in age, sex, education and time since diagnosis.^7^

If Parkinson’s disease with and without RBD are the same disease, but differ with respect to the time course of compensatory neuroplasticity (as reflected in measures of white matter FA), then it would be predicted that a progressive reduction in FA in the PD-RSWA group would eventually results in the development of RSWA in conjunction with worsening of motor and non-motor signs. This interpretation assumes that elevated corticofugal and corticocortical FA reflects compensatory processes that preserve motor and cognitive function and muscle atonia during REM sleep. However, the balance of evidence suggests that this is not the case. Studies that have estimated the prevalence of RBD and/or RSWA in PD have shown that a large proportion of people with Parkinson’s disease, including those with more advanced disease, do not have RBD or RSWA.^1-8^ Accordingly, we speculate that the majority of individuals in our PD-RSWA group will not develop RSWA or RBD, but motor and cognitive impairment will continue to progress along with decreasing FA. This hypothesis implies that the alterations in corticofugal and corticocortical white matter FA observed in this study impact motor and cognitive function, but not the REM sleep atonia circuitry. This interpretation is consistent with the idea that Parkinson’s disease with RBD and Parkinson’s disease without RBD are different diseases with respect to the topography and time course on non-dopaminergic alpha-synuclein pathology.^53^ There is growing evidence that people that co-express Parkinson’s disease and RBD have significant reductions in enteric, parasympathetic and sympathetic function, and increased involvement of brainstem nuclei and cortical structures. This is consistent with a more diffuse caudal-to-rostral distribution of synucleinopathy compared to those without RBD, despite similar levels of nigrostriatal dopaminergic denervation.^52, 54^ Accordingly, differences in FA, MD and RD between groups could be due to phenotypic differences in the characteristics of axonal microstructure (i.e. PD-RSWA have inherently higher FA), the spatial distribution of synuclein-related degeneration that impacts the integrity of axonal pathways, or the capacity for adaptive changes in structural and functional connectivity in response to nigral and extranigral neurodegeneration.

Increased white matter FA in early-stage Parkinson’s disease has been interpreted to reflect adaptive compensatory neuroplastic changes in structural connectivity.^17, 18, 28^ Increased FA could result from compensatory axonal sprouting in response to degeneration or alterations in the activity and/or synaptic connectivity of extranigral circuitry. Spontaneous behavioral recovery following partial lesions of the substantia nigra pars compacta in animal models of parkinsonism is associated with sprouting of collateral axons of surviving dopaminergic neurons.^55^ Adaptative and maladaptive alterations in the morphology and synaptic connectivity of extranigral pathways also occur following dopaminergic denervation.^16, 56-58^ Accordingly, abnormally elevated FA in people with Parkinson’s disease relative to controls may reflect compensatory neuroplastic changes in the microstructure of axons aimed at maintaining the function of extranigral pathways. Alternatively, higher white matter FA could also reflect degenerative processes that affect diffusion measures. Axonal degeneration is typically associated with reduced FA and increased in RD, MD and AD, reflecting decreased directionality of water diffusion and increased water diffusion respectively.^59, 60^ When imaging white matter, fibers from multiple directions can contribute to the FA value of a given voxel, therefore, making the average direction of diffusion of water more isotropic and yielding a lower FA value. If selective crossing fibers within the voxel degenerate, the direction that water diffuses will be more “unidirectional” or more anisotropic leading to higher FA. There is extensive fiber crossing in many of the pathways that showed significantly higher FA in the PD-RSWA group, such as the anterior, superior and posterior corona radiata.^61-63^ This raises the possibility that the higher FA observed in many of the white matter regions observed in this study reflect the loss of cortico-cortical or transcallosal crossing fibers that intersect corticofugal pathways.

Support for the idea that an increase in FA reflects a compensatory-like alteration in white matter microstructure comes from our data showing significant relationships between quantitative motor and cognitive measures, and diffusion measures in specific white matter tracts. Across all significant correlations, better motor and cognitive performance in the Parkinson’s disease group was associated with higher FA and lower RD and MD. Regression analyses further showed that performance in specific motor domains could be predicted by diffusion measures in specific pathways. The spatiotemporal measures of gait (normalized speed and step length) were predicted by FA of the anterior corona radiata. The anterior corona radiata include corticofugal projections from the prefrontal cortex that are considered to play a role in the cognitive control of gait.^64^ It has been hypothesized that increased activity in these pathways helps to compensate for the loss of gait automaticity associated with progressive degeneration of nigrostriatal and pedunculopontine neurons in people with Parkinson’s disease. If higher FA in prefrontal projection pathways in the PD-RSWA group reflects a compensatory adaptation in connectivity, this may explain the absence of a significant deterioration in gait, relative to healthy older adults, and the “unmasking” of gait impairment in PD+RSWA. Arm bradykinesia, as measured by the peak velocity of ballistic elbow movements, was predicted by FA of the superior corona radiata. Force generation mediating this task is largely controlled by the output from the motor and premotor cortices via the corticospinal tract.^65^ The gain in motor response is also dependent upon activity in the basal ganglia-thalamo-motor cortical pathway.^66, 67^ Increased inhibition of the thalamus in the parkinsonian state could be compensated for by increased connectivity of thalamocortical connections and excitability of motor cortical output pathways. Consistent with this idea, long-term potentiation (LTP)-like motor cortical plasticity, as measured using the paired-associative stimulation technique, is increased contralateral to the clinically less-affected arm and decreased contralateral to the clinically more-affected arm in people with untreated (de novo) Parkinson’s disease.^68^ These mechanisms could explain the retention of movement speed in the PD-RSWA group and emergence of bradykinesia in people with PD+RSWA.

Cognitive impairment is common in Parkinson’s disease, particularly in executive function and visuospatial domains.^69, 70^ Cognitive performance was significantly better in the PD-RSWA compared with the PD+RSWA group in measures of spatial-sequential working memory, visuospatial memory, and frontal executive function. This finding is consistent with studies showing that people with Parkinson’s disease and RBD are more likely to have impairment in executive function, attention and episodic learning and memory and visuospatial abilities compared to those without RBD.^14, 71^ Similar to the motor data, better performance in these cognitive domains was associated with higher FA and lower RD and MD. Visuospatial memory on the BVMT was significantly predicted by RD of the left posterior corona radiata. Perseverations during the card sorting task (a measure of executive function) was predicted by RD in the left anterior limb of the internal capsule. Zheng et al.^72^ have previously reported a similar relationship between diffusion measures in specific white matter tracts and cognitive performance in people with Parkinson’s disease, that is, higher FA (lower MD) was associated with better scores on tests of executive function, attention, memory and language and attention. For example, executive function scores inversely correlated with MD in the anterior and posterior corona radiata, anterior limb of the internal capsule, genu of the corpus callosum, and superior and inferior fronto-occipital fasciculi.

There are several technical features of the present work that may have contributed to differences in the findings relative to previous diffusion MRI studies in early Parkinson’s disease. We used ultra-high field MRI at 7T, which provides higher signal-to-noise and increased spatial resolution (1.25mm isotropic voxel size), relative to conventional imaging at 1.5T or 3.0T, to better capture the diffusion characteristics of individual pathways. The TBSS analysis used improved tensor-based registration ^39^ and spatial normalization to a template image created from our data. This has been shown to produce fewer false positives. DTI-TK has been shown to reduce misalignments of tracts, producing fewer false positives compared to the standard registration available in FSL, in a study conducted with synthetic FA maps with known FA values to represent the two groups.^39^ Care was taken to minimize movement artifacts by using MRI acquisition protocols that were optimized for the study of patients with Parkinson’s disease.^35, 73-75^ All testing was conducted in the practically-defined off-medication state to lessen the potential confound of the effects of dopaminergic medication on diffusion measures.^18^

It is important to reiterate that the Parkinson’s disease cohort tested in this study was separated post-hoc on the basis of measures of RSWA and not the presence/absence of dream enactment behavior or a confirmed diagnosis of RBD. Four of the 18 participants in PD+RSWA group did not report dream enactment or show complex movements or vocalizations during PSG testing. For this reason, our findings and conclusions are related to the expression of increased motor tone during REM sleep and not the presence/absence of RBD. Currently, we do not know if the abnormal elevation in corticofugal and corticocortical FA in the PD-RSWA group reflects an early compensatory mechanism that acts to preserve motor and cognitive function or reflects a characteristic of the PD-RSWA phenotype. In addition, the temporal dynamics of corticofugal and corticocortical FA in PD-RSWA vs. RD+RSWA are unknown. While the literature shows that group average FA decreases with increasing disease severity in PD,^19-27^ none of these studies separated the Parkinson’s disease cohort into those with and without RSWA or RBD. For this reason, we do not know if FA will continue to remain elevated in the PD-RSWA group, despite a progressive decline in motor and cognitive function over time, or decrease in concert with motor/cognitive signs. Prospective studies with long-term follow-up are needed to answer these questions.

## Supporting information

Supplement 1 - Methods for designation of RSWA groups

## Abbreviations

7T: 7 Tesla
AD: axial diffusivity
BDI: Beck Depression Inventory
BVMT: Brief Visuospatial Memory Test
D-KEFS: Delis-Kaplan Executive Function System
dMRI: diffusion magnetic resonance imaging
DRS: Dementia Rating Scale
FA: fractional anisotropy
MD: mean diffusivity
MDS-UPDRS: Movement Disorders Society Unified Parkinson’s Disease Rating Scale
MoCA: Montreal Cognitive Assessment
PSG: polysomnography
RBD: REM sleep Behavior Disorder
RD: radial diffusivity
REM: Rapid Eye Movement
RSWA: REM sleep without atonia
TBSS: tract-based spatial statistics
WAIS-IV: Wechsler Adult Intelligence Scale – IV
WCST: Wisconsin Card Sorting Test.

## Acknowledgements

We thank the volunteers that participated in this study. We are grateful for our additional referring movement disorders specialists including Dr. Martha Nance, MD, Dr. Daniel Kuyper, MD, and Dr. Sotirios Parashos, MD at Struthers Parkinson’s Center and Dr. Julia Johnson, MD at HealthPartners Parkinson’s Center.

