## Supplement 1 - Methods for designation of RSWA groups for "White matter microstructure in Parkinson’s disease with and without elevated REM sleep muscle tone"

**Classification of PD participants according to RSWA**

Individuals were recruited on the basis of a diagnosis of PD by the referring movement disorders neurologists according to the Movement Disorder Society clinical diagnostic criteria.^1^ Inclusion criteria included age 21-79 years and the ability to ambulate 50 m without an assistive device. Participants were excluded for any additional neurological disorder, a score of less than 22/30 on the Montreal Cognitive Assessment (MoCA),^2, 3^ musculoskeletal disorders that significantly affected movement, implanted deep brain stimulators or other surgeries to treat Parkinson’s Disease, pregnancy, untreated sleep apnea, or the presence of non-MRI compatible medical implants or devices.

All participants completed an overnight PSG to evaluate sleep and quantify RSWA. PSGs were scored by a single rater (AV), blinded to disease state, using the American Academy of Sleep Medicine Manual for the Scoring of Sleep and Associated Events to identify excessive sustained (tonic) and transient (phasic) muscle activity during REM sleep.^4^ The level of tonic or phasic RSWA was used to separate the Parkinson’s disease participants into normal (PD-RSWA) or abnormally elevated (PD+RSWA) muscle tone groups. Participants were classified as having PD+RSWA based on the percentage of REM sleep with elevated tonic or phasic muscle activity in the chin (submentalis), arms (flexor digitorum superficialis) or legs (tibialis anterior).

All participants, including controls, completed an overnight polysomnography (PSG) study at the Sleep Center at the University of Minnesota, Fairview. Participants were allowed to take antiparkinson medications during the PSG. The sleep studies were performed using standard video-based PSG procedures, including electromyography (EMG) recordings from the submentalis (chin), flexor digitorum superficialis and tibialis anterior (TA) muscles as well as electroencephalography recordings from 10 scalp electrodes. PSG data was analyzed by a blinded rater who is board certified in sleep medicine (A.V.). Sleep stages and percentage of REM sleep with RSWA were scored according to the American Academy of Sleep Medicine Manual for the Scoring of Sleep (Berry et al., 2018).

Tonic and phasic muscle activity during REM sleep were quantified for the submentalis (chin) EMG signal and phasic activity was quantified for the TA muscle. Tonic muscle activity was defined as the presence of increased EMG activity during REM sleep exceeding twice the background activity in stage N3 sleep. Each 30-second epoch was scored as tonic or atonic depending on whether tonic chin EMG activity was present for >=50% or <50% of the epoch. The percentage of REM sleep with tonic muscle activity (% tonic EMG) was derived from the sum of all epochs containing increased tonic muscle activity divided by the REM sleep time. The phasic muscle activity was assessed in 3-second mini-epochs during REM sleep. A phasic EMG event was defined as any burst of EMG activity lasting 0.1 to 5 seconds with amplitude exceeding 4 times the background EMG activity. The percentage REM sleep with phasic muscle activity (% phasic EMG) was derived from the sum of all mini-epochs containing increased muscle phasic activity divided by the REM sleep time.

Participants with PD were stratified into low and high RSWA groups based on the percentage of time during REM sleep when RSWA was present. The threshold for separation into RSWA negative vs. RSWA positive groups was derived from the distributions of the RSWA PSG scores across all subjects, including the controls. Participants with RSWA above threshold in either the tonic or phasic chin or phasic leg muscles and/or clear evidence of dream enactment were assigned to the RSWA+ group (arm EMG was not used in the classification due to poor quality signals in many of the participants). All participants in the RSWA+ group had elevated phasic or tonic submentalis EMG and 14 of 18 (78%) had self-reported or partner-reported dream enactment and/or DEB was observed during the PSG.

Difference in RSWA across groups were analyzed using a univariate ANOVA with age and sex as covariates. There was a significant effect of group for the tonic and phasic chin, phasic leg, and combined phasic chin+leg RSWA scores (F (2) > 13.0, p < 0.001). Post-hoc comparisons using Tukey’s test showed that RSWA scores for all variables in the PD+RSWA group were significantly higher compared to the PD-RSWA group and control groups (p < 0.001). There was no significant difference in tonic and phasic chin, phasic leg, or chin+leg RSWA scores between the control and PD-RDWA groups (p > 0.688).

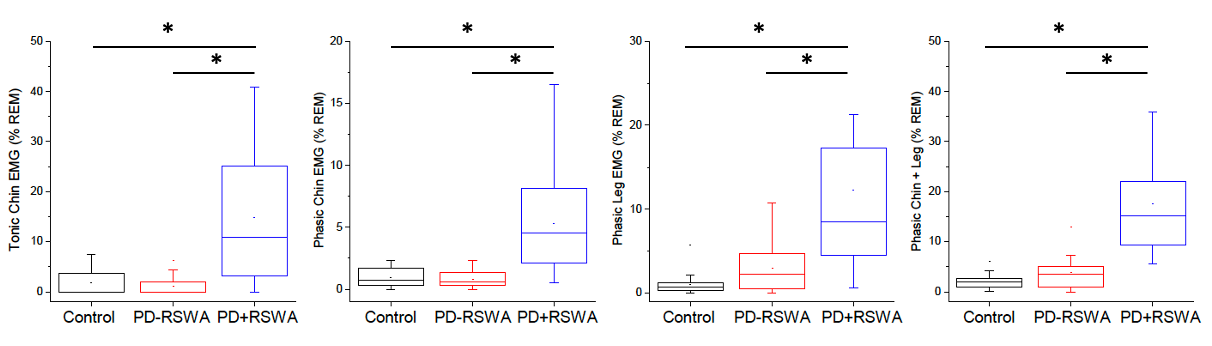

Supplement Figure. 1. Rapid eye movement (REM) sleep without atonia (RSWA) values for tonic and phasic submentalis (chin), phasic tibialis anterior (leg) and combined phasic chin and leg EMG across groups. There was a significantly higher level of RSWA in the PD+RSWA group compared to the PD-RSWA and control groups in all muscles or muscle combinations (one-way ANOVA, Tukey’s post hoc comparisons, * p < 0.001). Boxes range from the first to the third quartile, whiskers extend to 95% confidence intervals, median is indicated by a line across the box, and mean is indicated by a square marker.

*References:*

1.	Postuma RB, Berg D, Stern M, et al. MDS clinical diagnostic criteria for Parkinson's disease. Movement disorders. Oct 2015;30(12):1591-601. doi:10.1002/mds.26424

2.	Nasreddine ZS, Phillips NA, Bedirian V, et al. The Montreal Cognitive Assessment, MoCA: a brief screening tool for mild cognitive impairment. J Am Geriatr Soc. Apr 2005;53(4):695-9. doi:10.1111/j.1532-5415.2005.53221.x

3.	Karlawish J, Cary M, Moelter ST, et al. Cognitive impairment and PD patients' capacity to consent to research. Neurology. Aug 27 2013;81(9):801-7. doi:10.1212/WNL.0b013e3182a05ba5

4.	Berry RB, Brooks R, Gamaldo CE, Harding SM, Lloyd RM. AASM Manual for the Scoring of Sleep and Associated Events: Rules, Terminology, and Technical Specifications, Version 2.5. 2018.
